# Systematic Review of Monogenic Diabetes Prognostics

**DOI:** 10.1101/2023.05.19.23290220

**Authors:** Rochelle N. Naylor, Chloé Amouyal, Louis H. Philipson, Camille Vatier, Laura T. Dickens, ADA/EASD PMDI, Siri Atma W Greeley

## Abstract

**Background:** Individuals with monogenic diabetes are at risk for diabetes-related complications; however, overall prognosis and whether prognosis is similar to other diabetes forms is poorly understood.

**Aim:** To assess diabetes-related microvascular and macrovascular complications in the common forms of monogenic diabetes.

**Methods:** Systematic review with data sources from Pubmed, Medline and Embase was performed to assess diabetes-related complications in KCNJ11-neonatal diabetes, ABBC8-neontal diabetes, HNF1A-diabetes, HNF4-diabetes and GCK-related hyperglycemia.

**Results:** Data was extracted from 67 studies. Most studies had moderate to high risk of bias. In neonatal diabetes, 16 of 20 studies reported at least one microvascular complication, with complications occurring as early as the second decade of life. Macrovascular complications were reported in only 1 individual who was 40 years old at the time of study. Diabetes complications were frequent in HNF1A-diabetes and HNF4A-diabetes, but did show a temporal trend of improved prognosis (e.g., 47% versus 13.6% retinopathy) and better prognosis compared to type 1 diabetes. Death due to cardiovascular disease was higher in HNF1A-diabetes compared to unaffected relatives (66% versus 43%). GCK-related hyperglycemia showed overall low rates of complications.

**Conclusion:** While KCNJ11-neonatal diabetes, ABBC8-neontal diabetes, HNF1A-diabetes and HNF4-diabetes are clearly at risk for diabetes-related complications, microvascular complications were infrequently reported before the third decade of life. GCK-related hyperglycemia showed a low prevalence of complications with rates not significantly different from control groups except for mild retinopathy. Future prospective studies to determine age at onset of complications and the impact of precision therapy are warranted to best guide surveillance practices for each subtype.

## Introduction

Diabetes-related complications are a major source of healthcare expenditures, lost productivity and reduced quality of life resulting from the associated morbidity and earlier mortality. Applying a precision lens to prognosis for different types of diabetes will better inform surveillance and treatment recommendations and improve outcomes. Similar to polygenic forms of diabetes, individuals with monogenic diabetes are at risk for developing diabetes-related microvascular and macrovascular complications. There are some shared risk factors with both type 1 (early-onset) and type 2 diabetes (delayed recognition of diabetes, with some individuals already having complications at the time of diagnosis). However, there are also important differences in the common forms of monogenic diabetes that may have additional impact on prognosis and warrant systematic review to guide clinical practice.

Neonatal diabetes mellitus (NDM) occurring under 6-12 months of age occurs in about 1:100,000 births, with the most common cause being due to heterozygous activating mutations in the two genes – KCNJ11 and ABCC8 – encoding the beta-cell KATP channel.^1^ This lifetime of exposure to elevated blood glucose levels could result in excess and early morbidity, as suggested in studies of type 1 diabetes where 45% of children <5 years old at diabetes diagnosis experienced persistent microalbuminuria (a risk factor for nephropathy) before puberty onset compared to those diagnosed between 5-11 years who only experienced microalbuminuria at or after puberty.^2^ However, individuals carrying pathogenic variants in *KCNJ11* and *ABCC8* will usually respond well to treatment with oral sulfonylurea medications that represent a precision therapy, resulting in rapid and marked improvement in HbA1c without the marked fluctuations in blood glucose or significant rates of hypoglycemia seen in insulin therapy.^3^ This optimal glycemic control presumably leads to a reduction of risks of diabetes-related complications.^4^ In addition to the barriers of genetic diagnosis and implementation of appropriate therapy, understanding of long-term outcome currently relies on reports of cases and cohort studies limited by the rarity of these disorders and the relatively short period of time they have been known (since 2004).^5^

Heterozygous mutations in the HNF1A and HNF4A beta-cell transcription factors are among the most common causes of autosomal dominant, young-onset diabetes, also known as maturity-onset diabetes of the young (MODY). HNF1A-diabetes (also known as HNF1A-MODY or MODY3) and HNF4A-diabetes (HNF4A-MODY or MODY1) are characterized by young-onset diabetes (typically second and third decades of life) owing to a progressive insulin secretory defect with preserved insulin sensitivity. Rates of overweight and obesity are similar to the background population. These forms of diabetes also tend to be responsive to treatment with sulfonylureas, and demonstrate hypersensitivity with low doses often sufficient to achieve good glycemic control. However, similar to type 2 diabetes, individuals with HNF1A-diabetes and HNF4A-diabetes may have a substantial lag in diagnosis after onset of diabetes and present with complications at the time of diagnosis, which may exaggerate the apparent risk for diabetes complications. The autosomal dominant inheritance of HNF1A-diabetes and HNF4A-diabetes typically means that subsequent generations are diagnosed earlier and temporal trends in time to diagnosis and time to precision treatment between older and younger generations may be able to better inform overall prognosis for these forms of diabetes as well as the ideal age range or duration after diabetes onset to begin surveillance for complications. In HNF1A-diabetes, the genetic impact of pathogenic *HNF1A* variants on traditional risk factors for cardiovascular disease may also mean that guidelines specific to HNF1A-diabetes may be warranted.

GCK-related hyperglycemia (also known as GCK-MODY or MODY2) has a unique phenotype of lifelong, mild hyperglycemia with HbA1c typically ranging from 5.6-7.6%. For many individuals, their HbA1c will remain within the HbA1c goal range used for most non-pregnant adults of <7%^6^ without therapy. Even for individuals whose HbA1c falls in the range of >7.0-7.6%, pharmacologic treatment is not advised and the body of literature supports that HbA1c is not altered by glucose-lowering therapy in isolated GCK-related hyperglycemia.^7^ Reported rates of diabetes-related complications are low but not absent. Except for background retinopathy, it is not clear that rates of microvascular or macrovascular complications differ from the population of people who do not have diabetes. It is also not clear what impact obesity has on prognosis in GCK-related hyperglycemia or how to distinguish between the elevated HbA1c owing to heterozygous pathogenic variants in the *GCK* gene and the dysglycemia of type 2 diabetes.

Systematic review of diabetes-related microvascular and macrovascular complications in monogenic diabetes will allow a summation and appraisal of the strength of the published literature on monogenic diabetes prognosis in order to optimize screening practices and ultimately improve outcomes and quality of life in those living with monogenic diabetes.

## Aims

We aimed to provide a systematic review of the evidence for monogenic diabetes prognostics for the microvascular and macrovascular complications in patients with neonatal diabetes due to mutations in*KCNJ11* and *ABCC8* and in patients with autosomal dominant young-onset diabetes (also known as maturity-onset diabetes of the young, MODY) due to mutations in *HNF1A*, *HNF4A* and *GCK.* We had initially hoped to address severity as well but due to inconsistencies in how complications were reported, we mainly focus on their presence or absence in case reports and case series and their prevalence in cohort studies.

## Methods

We registered the protocol for this systematic review in Prospero (https://www.crd.york.ac.uk/prospero/: CRD42023399458.

Data Sources: We searched available literature on monogenic diabetes prognostics in PubMed, MEDLINE, and Embase. Our full search strategy is available in Supplemental Table 1.

Study Selection: We utilized Covidence (https://www.covidence.org) to carry out filtering and selection of studies. Title and abstract review was carried out by four authors with each title and abstract reviewed by two authors and a third author resolved discrepancies. Full text review was carried out by all authors, with each text receiving two votes with discrepancies resolved by a third review. Each author contributed to data extraction using a standardized tool. Data extracted included publication year, study design, number of participants and participant information including sex, age at diabetes diagnosis and at time of study, glycemic data, and diabetes-related complications.

Inclusion and Exclusion Criteria: We included original English language, human studies published after 1992 that had full text available and that provided information on microvascular and/or macrovascular complications for the following genes and phenotypes:

Permanent neonatal diabetes due to pathogenic variants in *KCNJ11* and *ABCC8*
Autosomal dominant, young onset diabetes (MODY) phenotype due to pathogenic variants in*GCK*, *HNF1A*, and *HNF4A*.

We excluded studies or data within studies of individuals with non-pathogenic variants, with co-occurrence of type 1 diabetes or of two types of monogenic diabetes, reporting on a phenotype outside of the scope of this review (*KCNJ11* or *ABCC8* mutations resulting in transient neonatal diabetes or MODY phenotypes or*HNF1A*, *HNF4A*, or *GCK* homozygous mutations resulting in neonatal diabetes) or studies that did not report on the outcomes of interest.

Quality assessment: We used the methods outlined in Murad et al.^8^ and the study quality assessment tools available from NHLBI to assess risk of bias for extracted studies (https://www.nhlbi.nih.gov/health-topics/study-quality-assessment-tools).

Data synthesis: Data are presented as single subject data or as mean ± standard deviation, median [interquartile range, IQR], median (range) or as percentages. We did not carry out meta-analysis as there was significant heterogeneity in how diabetes-related complications were assessed and reported.

## Results

### Quality assessment

All studies had moderate or high risk of bias. Studies consisted of case reports, case series or cohorts. There were no controlled studies. For cohort studies, the recruitment sources often differed between groups. Many cases were recruited from monogenic diabetes registries while control groups were recruited from clinics or were unaffected family members. As control groups for monogenic diabetes were either other forms of monogenic diabetes, unaffected individuals, people with type 1 or type 2 diabetes, there were important differences between groups (e.g., either age at time of study or diabetes duration and BMI in studies comparing monogenic diabetes and type 2 diabetes). Not all studies reported how complications were assessed. In those studies with a specific aim to assess diabetes-related complications, investigators assessing complications were not always blinded to group assignments. In many case reports or case series, it was not clear if the absence of a reported diabetes-related complication was because it was not present in the individual or it was not assessed.

- **Question:** What are the risks and severity of diabetes-related microvascular and macrovascular complications in patients with permanent neonatal diabetes due to pathogenic variants in*KCNJ11* and *ABCC8* and in patients with maturity-onset diabetes of the young (MODY) due to pathogenic variants in *HNF1A*, *HNF4A* and *GCK*?

From the literature search, we identified 1747 studies. There was manual and automated exclusion of 296 duplicates. Of the remaining studies, 367 were selected for full text review after review of titles and abstracts. Data was extracted from 67 studies, including 12 studies that reported on multiple gene causes with the remaining reporting on one of the five monogenic diabetes subtypes of interest (Figure 1). Of the 67 studies, 18 studies contributed data on KCNJ11-neonatal diabetes and 3 studies contributed data on ABCC8-neonatal diabetes, 34 studies contributed data on HNF1A-diabetes, 6 studies on HNF4A-diabetes, and 19 studies on GCK-related hyperglycemia (Table 1). Where cohort studies were likely to represent significant overlapping patient data, presented data was taken from the most recent publication(s) to capture all relevant complications data.

**Figure 1.**
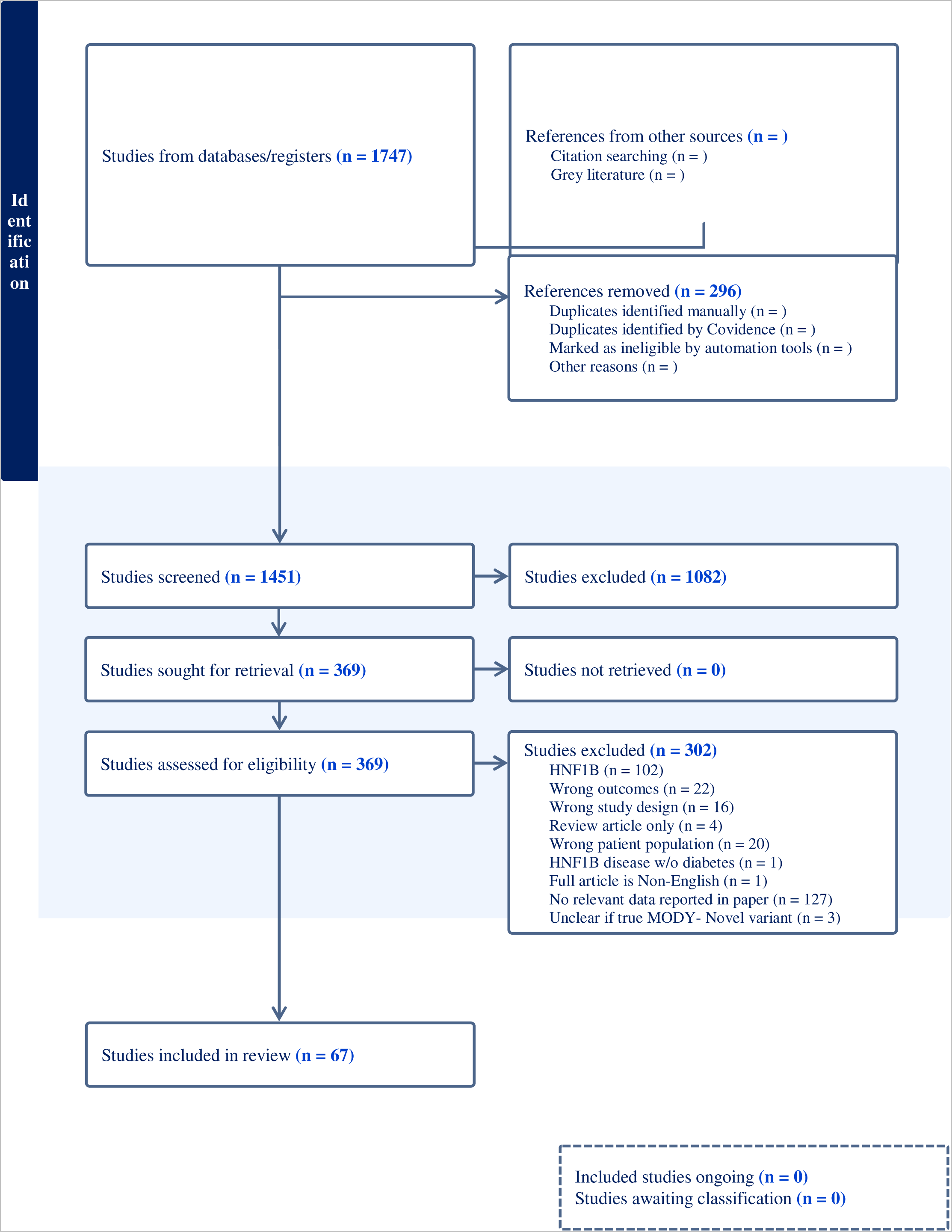
Schematic of selection of studies.

**Table 1.**
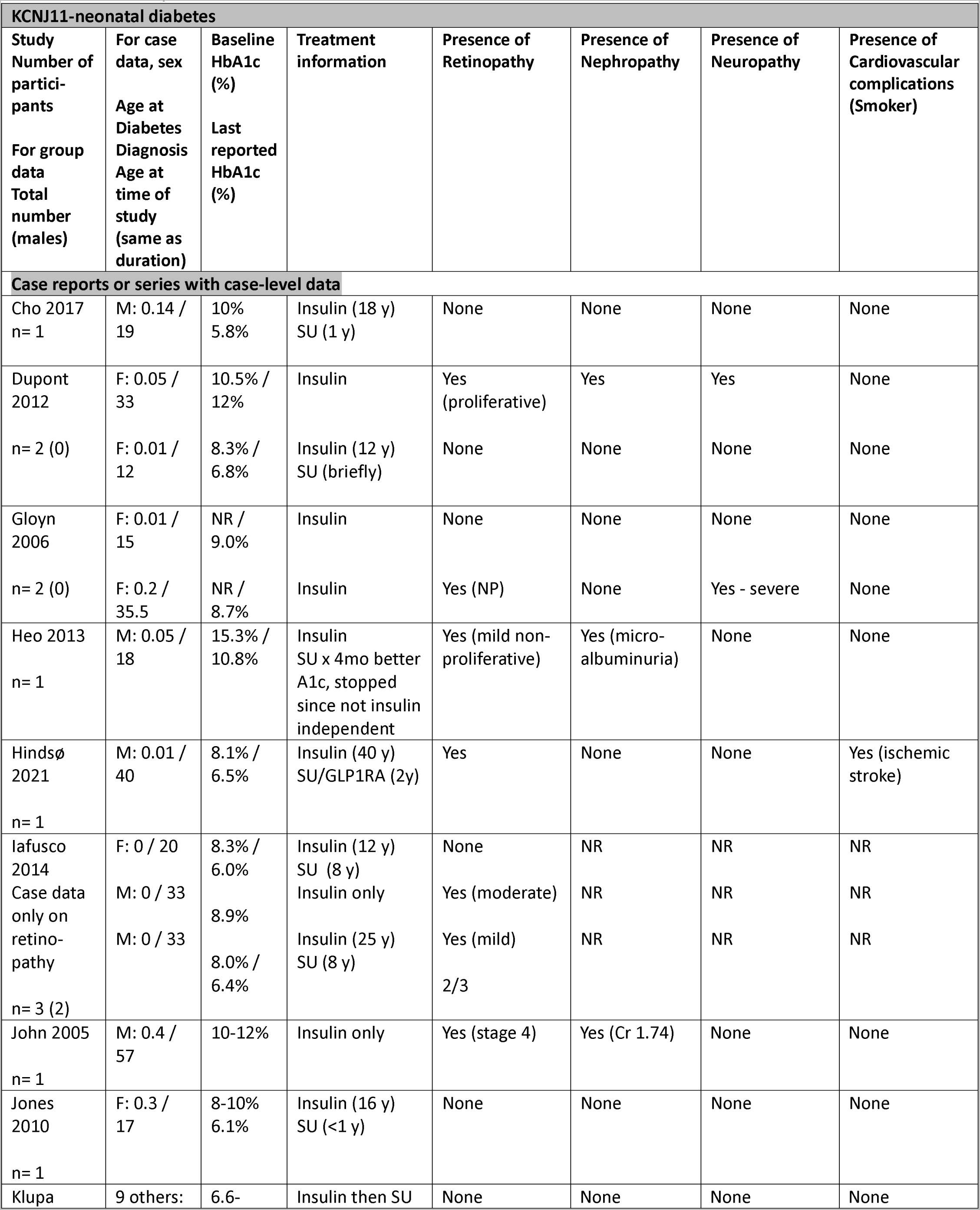
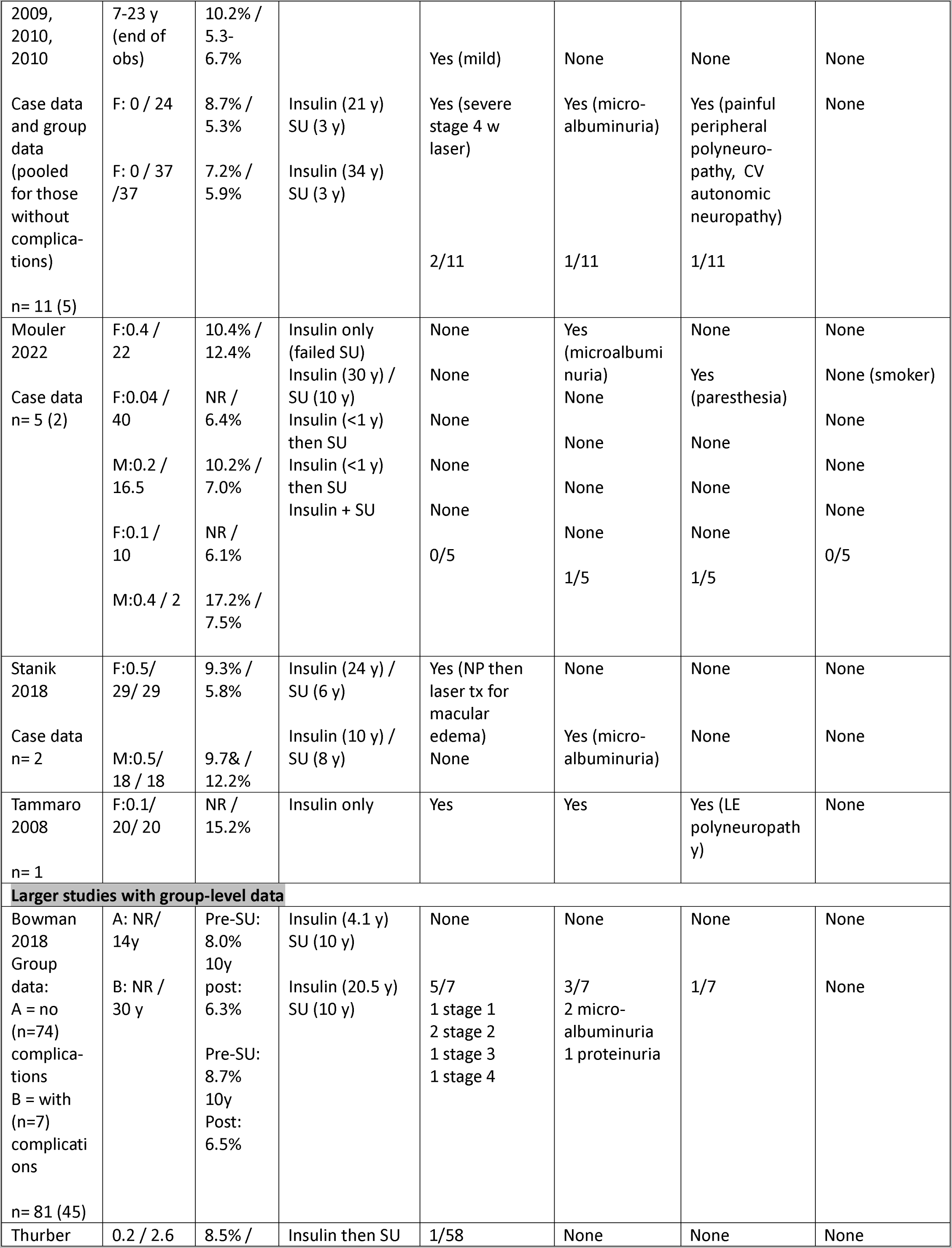
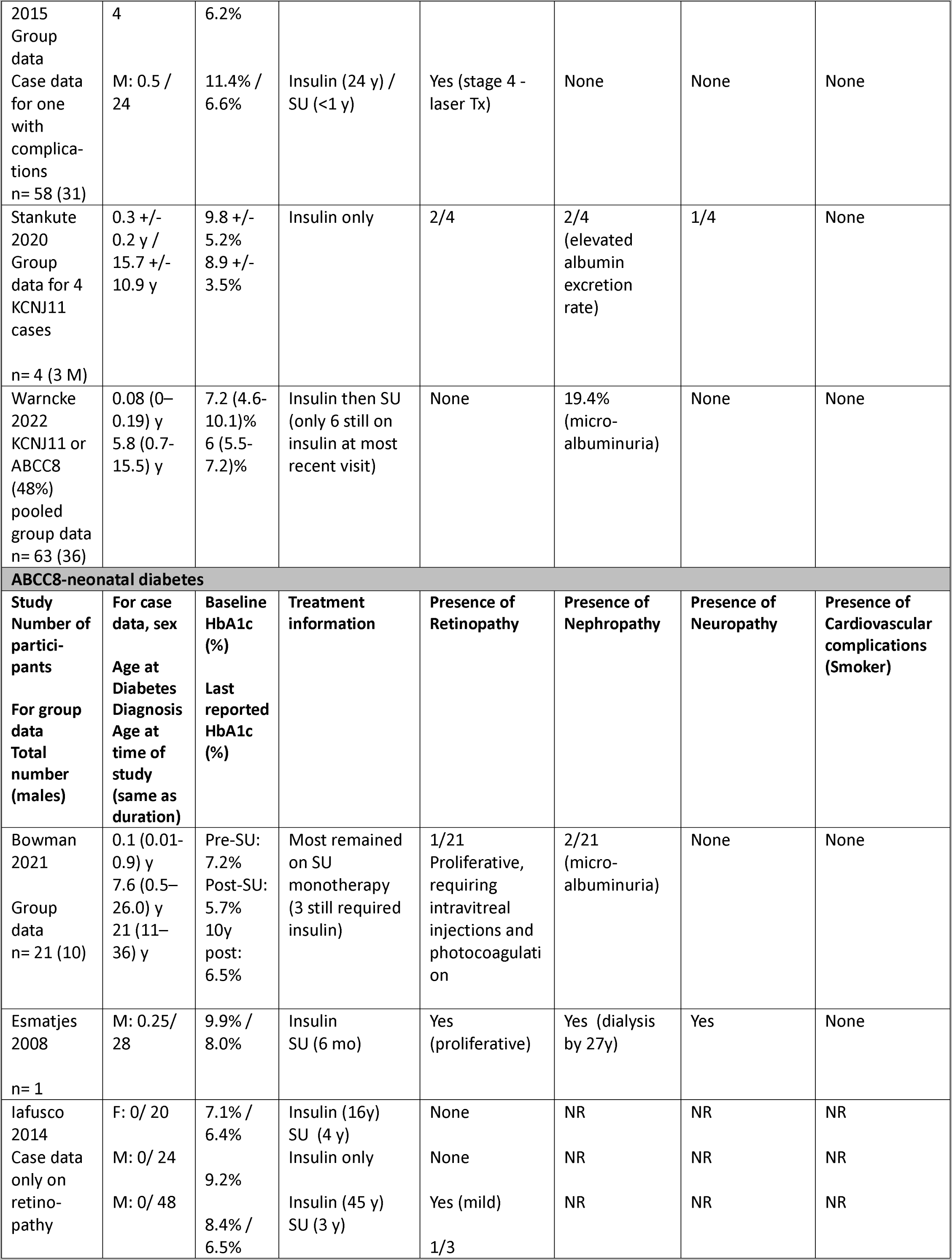
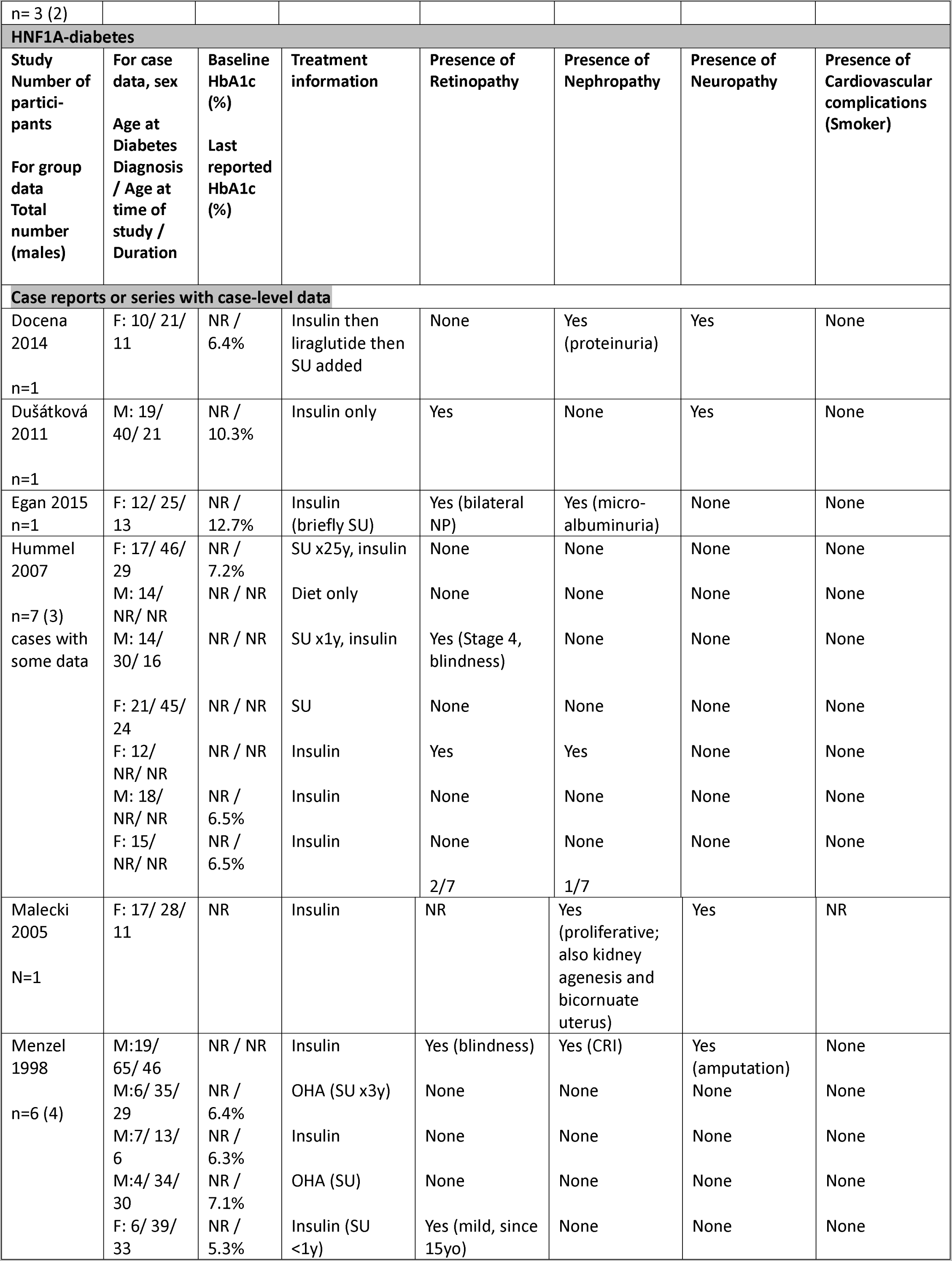
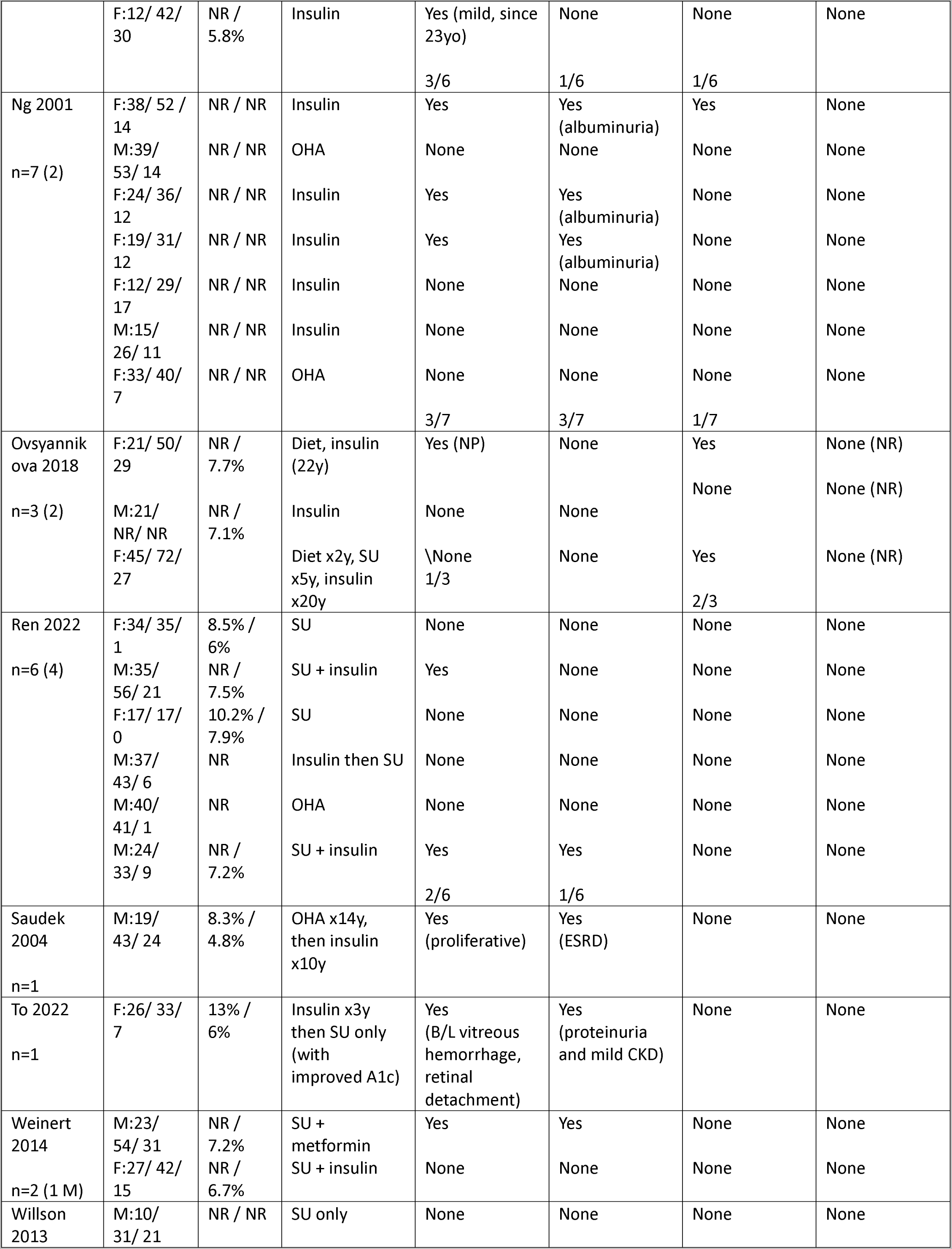
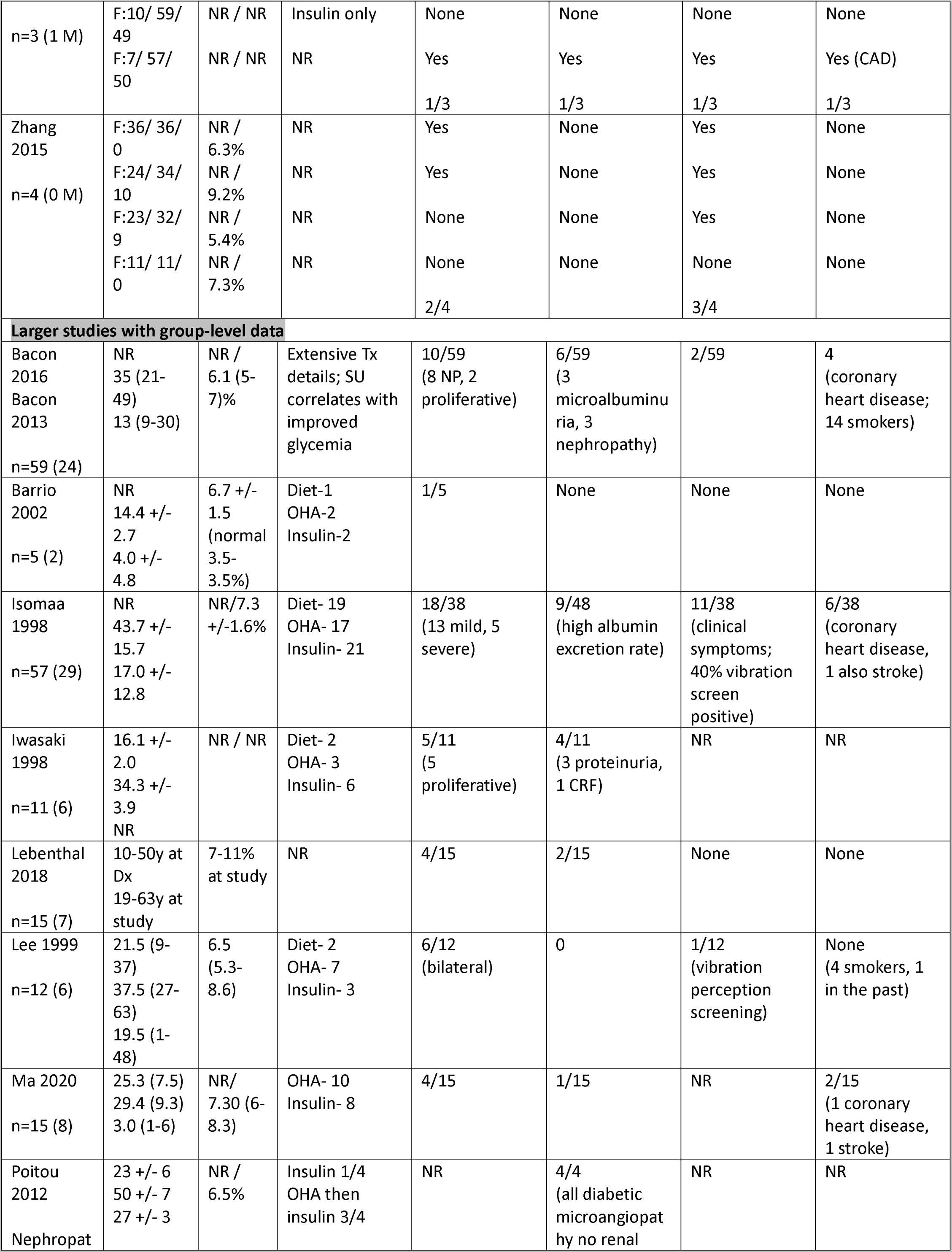
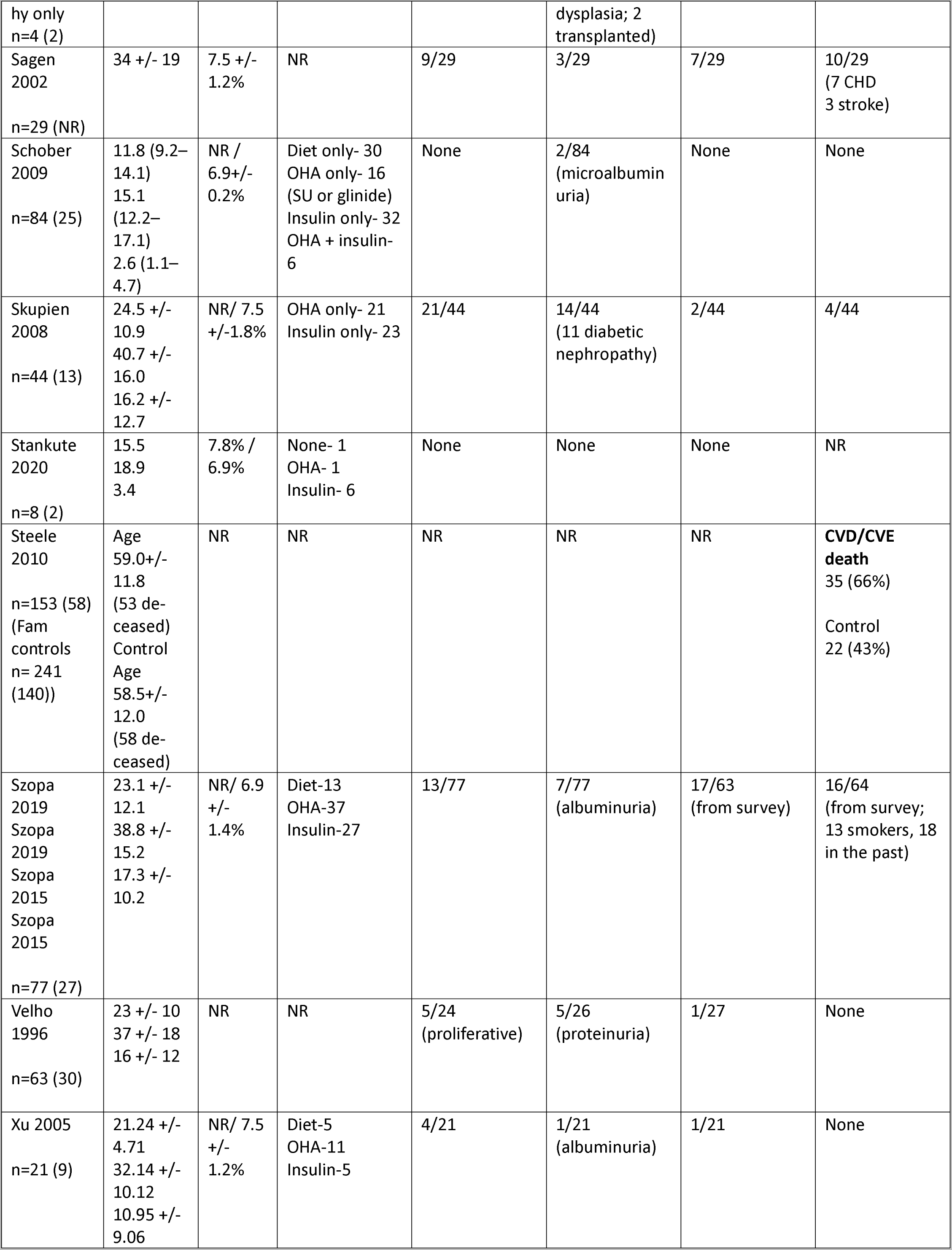
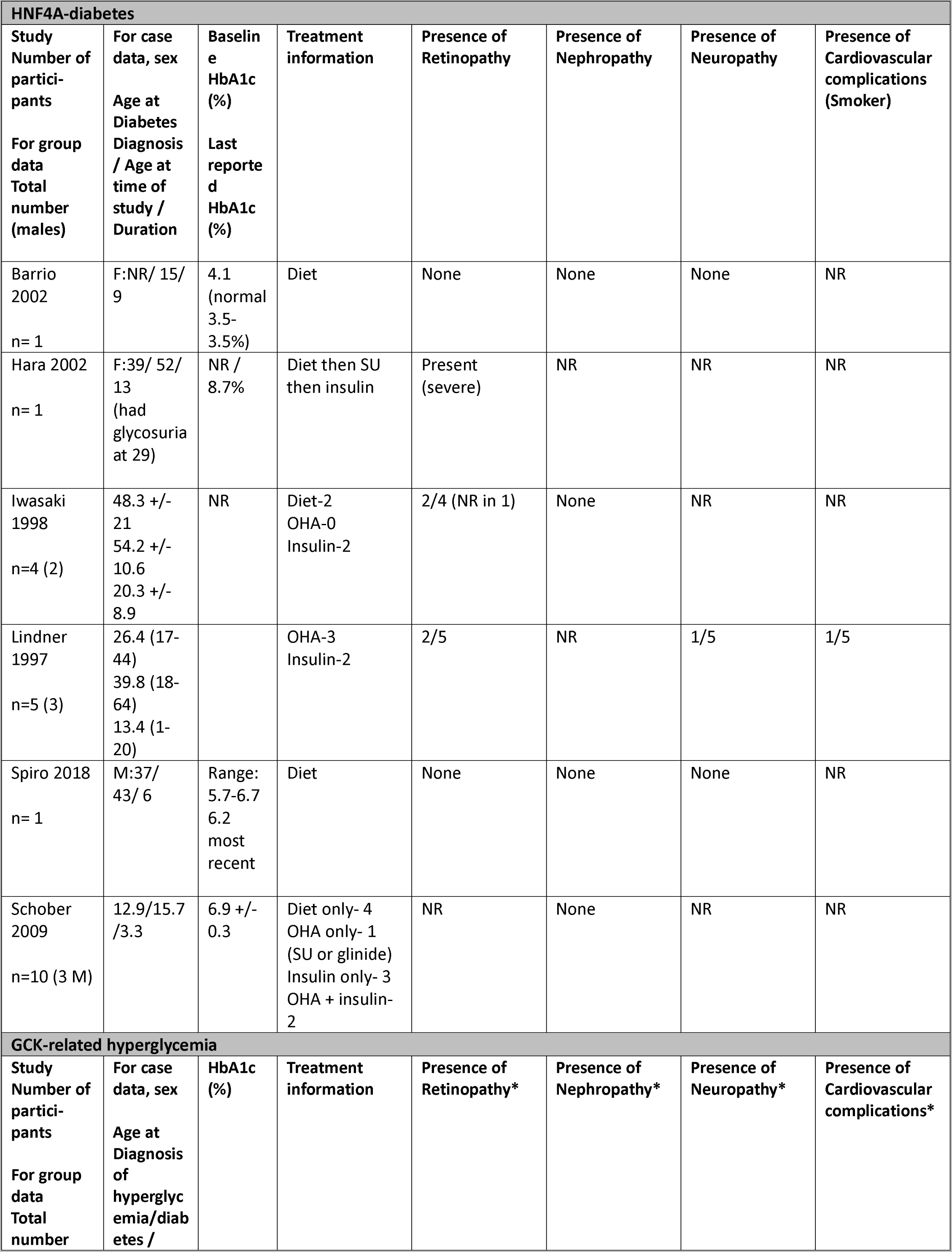
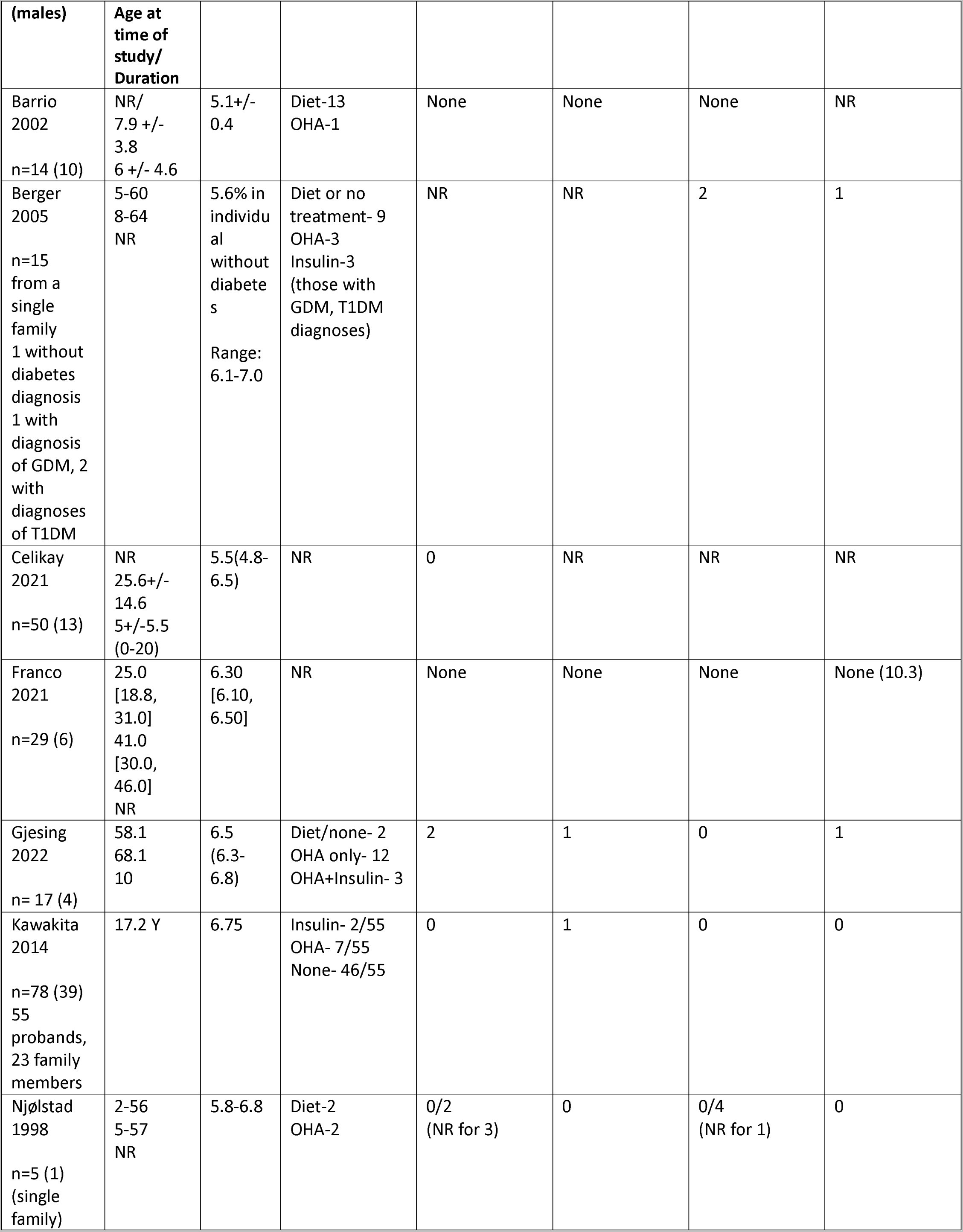
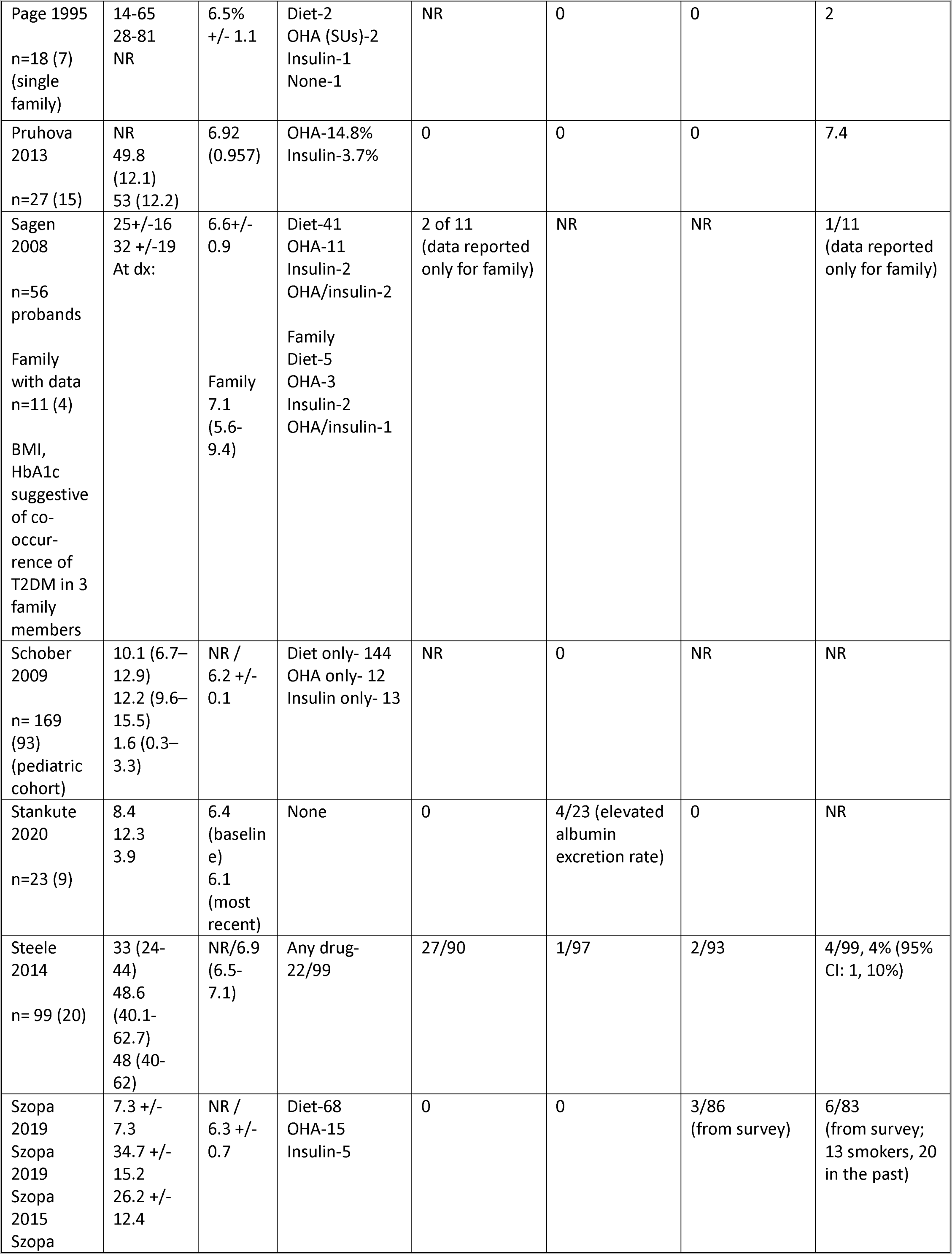
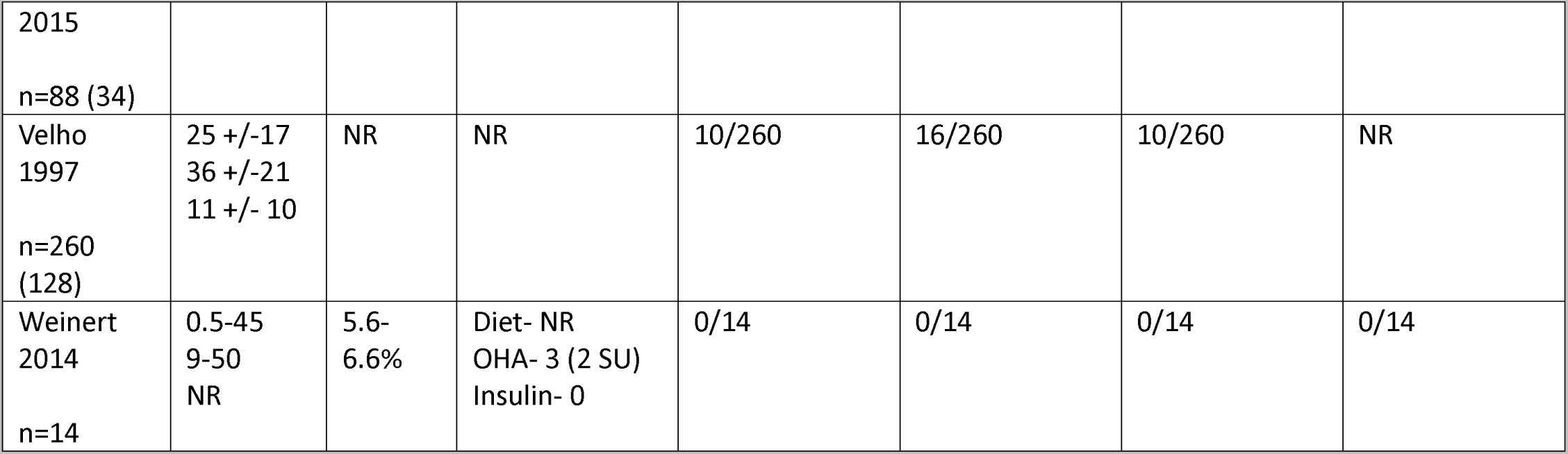
Summary of included studies.

### KCNJ11 and ABCC8 permanent neonatal diabetes

We identified 20 studies with data on diabetes complications in individuals with either KCNJ11 or ABCC8 related permanent neonatal diabetes. We extracted case-level data from 14 studies reporting on 31 individuals with KCNJ11-neonatal diabetes,^9–22^ as well as pooled group data from 4 studies representing 206 individuals.^23–26^ For ABCC8-neonatal diabetes, there was case-level data on 4 individuals and pooled group data on 21 individuals from 3 studies.^14, 27, 28^ Age at the time of study from the larger cohort studies ranged from 2.64-30 years in KCNJ11-neonatal diabetes and 20-48 in ABCC8-neonatal diabetes. Within case reports, age ranged from 2-57 years and 20-48 years in KCNJ11-neonatal diabetes and ABCC8-neonatal diabetes, respectively. The youngest age with complications (19.4% with microalbuminuria) reported was in a cohort with mean age of 5.8 years but range up to 15.5 years.^26^ There was also a case report of microalbuminuria and mild non-proliferative retinopathy in an 18-year-old with uncontrolled diabetes. Most complications occurred in the third decade of life or later. There was only 1 reported macrovascular complication of ischemic stroke occurring in a 40-year old male.^13^

### HNF1A-diabetes and HNF4A-diabetes

There was case level data for 44 individuals with HNF1A-diabetes from 14 studies.^29–42^ At least one individual had a microvascular complication in all 14 studies. The youngest age at time of study with a complication (proteinuria) was 21 years. There was also a 39 year-old female who was reported to have onset of mild retinopathy at 15 years of age (after 9 years of diabetes). In cases with complications where medication details were reported, all but one were receiving insulin as monotherapy or combination therapy. Only approximately 30% of individuals were using sulfonylureas (precision therapy) with some details in treatment data being difficult to clearly interpret. In the grouped data (n=657) both microvascular and macrovascular complications were common.^25, 43–61^ Moreover, Steele et al., showed that prevalence of cardiovascular disease was higher and related prognosis was worst in those with HNF1A-diabetes compared to their unaffected relatives.^55^ Similar to the case level data, there was complications occurring as early as the second decade of life-occurrence of retinopathy in a cohort with mean age 14.4 years and rare microalbuminuria in a cohort with mean age 15.1 years. Of note, comparing complication rates across older to newer studies, there is evidence for a temporal trend of improving prognosis. Isomaa et al.’s 1998 study of complications showed mean, SD HbA1c of 7.3% ± 1.6 with 33% on no pharmacologic therapy, 30% on oral agents, and 37% on insulin.^46^ In this cohort, 34% had mild retinopathy, 13% had severe retinopathy, 19% had increased albumin excretion rate, 29% had clinical neuropathy, and 16% had coronary heart disease (CHD). The rates did not statistically differ from “ insulin dependent” and “non-insulin dependent” diabetes, except for CHD, which was more prevalent in HNF1A-diabetes compared to insulin dependent diabetes but lower than in non-insulin dependent. In Bacon et al.’s 2016 study, median HbA1c was 6.1%, IQR [5-7]. There was active transition to sulfonylureas, with 32 of the 39 people not already on sulfonylureas transitioned to sulfonylurea therapy with decrement in HbA1c (60% of those on sulfonylureas maintained this therapy over 84 months of follow-up). When looking at complications, 13.6% had retinopathy with 70% having background retinopathy. Only 5% had persistent microalbuminuria, 5% had overt nephropathy, 3.3% had neuropathy, and 6.7% had CHD. These complication rates were overall lower than a cohort of type 1 diabetes.

Studies of HNF4A-diabetes represented a total of only 22 patients.^45, 47, 53, 62–64^ Five individuals (23% of all reported cases) had retinopathy. This included one severe case with near-blindness where there was evidence of a 10-year delay in diabetes diagnosis (Hara). In another case from a larger study of multiple forms of monogenic diabetes, there were only three individuals with HNF4A-diabetes. The mean age at diabetes onset was 5.8 years with a range of 1.9-13.3 years and all three had ketosis at diabetes and were treated with insulin. Thus, the possibility of co-occurring type 1 diabetes could be possible, but data was included as there was not a clear co-diagnosis of type 1 diabetes. Only one individual had renal abnormality (elevated albumin excretion rate) and two had neuropathy. Only one study reported on macrovascular complications with one person affected.^63^ Mean diabetes duration was over a decade in all cases of complications.

### GCK-related hyperglycemia

Rates of any complications were low in all studies of GCK-related hyperglycemia, but all types of microvascular complications were reported.^25, 40, 45, 53, 56–59, 65–75^ In case-level data, there was evidence of co-occurrences of type 2 diabetes (obese BMI and HbA1c outside of the expected range of GCK-related hyperglycemia), such that complications could not always be definitively attributed to carrying a pathogenic*GCK* variant. In the larger studies, no complications were reported in a pediatric cohort of 169 patients with median age of 12.2 years [IQR 9.6-15.1] at the time of study^53^, but another pediatric cohort (mean age, SD 12.3 ± 5.8 years at time of study) reported 17.4% prevalence of microalbuminuria.^25^ However, this study did not evaluate for persistent microalbuminuria. While both persistent and intermittent microalbuminuria are risk factors for diabetic nephropathy, intermittent microalbuminuria confers a much lesser risk.^76^ Notably in one pediatric and adult cohort (mean age, SD at time of study 36 ± 21 years), prevalence of kidney disease (proteinuria) was no higher than 6% (Velho 1997). Prevalence of proliferative retinopathy in this same cohort was 4%. In another study of 99 individuals with GCK-related hyperglycemia (median age of 48.6 years [IQR 40.1-62.7]), overall prevalence of diabetes microvascular and macrovascular complications were low and not significantly different from their unaffected relatives (controls) and were substantially lower than in type 2 diabetes.^74^ The most notable finding was that 30% of those with GCK-related hyperglycemia had background retinopathy (not requiring treatment), which was significantly lower than rates in type 2 diabetes (63% with 28% requiring laser treatment), but higher than controls (14%). Severity of retinopathy was less than seen in type 2 diabetes, not requiring treatment compared to 28% of those with type 2 diabetes receiving laser treatment for retinopathy. Prevalence of macrovascular complications was similarly not significantly different from their unaffected relatives.

## Discussion

To our knowledge, this is the first systematic review of diabetes prognosis in common forms of monogenic diabetes. Due to several distinct features of monogenic diabetes it is important to understand how diabetes prognosis may differ from type 1 and type 2 diabetes. These distinct features include the essentially lifelong presence of diabetes in neonatal forms, which may translate to earlier onset of complications. There is also the lifelong presence of mild hyperglycemia in heterozygous*GCK* mutations that is not altered by pharmacologic therapy but where many affected individuals will already have HbA1c at goal range as defined by the ADA.^6^ There is additionally the potential of earlier recognition and diagnosis in autosomal disease and the availability of precision therapy (sulfonylureas) relevant for HNF1A-diabetes, HNF4A-diabetes, KCNJ11-diabetes and ABCC8-diabetes, where risk for microvascular and macrovascular complications will be impacted by early and sustained good glycemic control. We have reviewed the evidence available to better understand diabetes prognosis in these forms of monogenic diabetes.

There are a number of limitations to the current data on diabetes prognosis in the common forms of monogenic diabetes. As is true in most rare conditions, a high risk of bias is present in nearly all of the described work. Thus, we did not use bias to exclude studies. Another significant limitation of the data is that most cases report complication status at a given timepoint, and do not report the age when the complication was first diagnosed. Moreover, in many studies the presence of certain complications was reported but it was often not clear if other complications had been screened for and were absent. Thus, it is unknown whether prevalence of complications are being over- or underestimated in these studies. There are exceptions to this, such as the German DPV registry that collects data on all possible complications on an ongoing basis in a more systematic manner.^26^ This kind of effort will be needed to gather detailed information over decades of life to better delineate risks and best treatment approaches. There is also inconsistency in the specificity of complications, limiting ability to assess severity. Treatment for complication risk factors, such as statin therapy or anti-hypertensive treatment for CV risk factors wasn’t accounted for as many studies either did not report on this or those treatment and those experiencing CV outcomes could not be linked.

Finally, while we expect the improved glycemia associated with precision therapy to translate into improved diabetes prognosis, studies were not designed to allow for comparison of complication outcomes based on precision and non-precision treatment.

### Recommendations for surveillance for KCNJ11 and ABCC8 permanent neonatal diabetes

While it is very clear that KATP individuals are at risk for the known glycemia-related diabetes complications (especially microvascular, but also some evidence of macrovascular), the data is not complete enough to analyze (e.g., by years of exposure to higher HbA1c levels) but there is some evidence that the complications are related to higher levels of glycemia (e.g., a case report might state that individual has had “poor DM control”) as for other forms of diabetes.

For KATP-related NDM, there is strong evidence for reduction of glycemia with SU treatment, but it remains uncertain to what extent initiation of SU will prevent DM complications, especially when started later in life after years of exposure to high levels of glycemia. Similarly, a few of the reports reviewed here noted that complications such as retinopathy had already developed prior to switching from insulin therapy to SU. The limited evidence in these few reports suggests that the complications did not worsen after SU treatment, but studies of larger numbers of patients of greater periods of time will be needed to strengthen the evidence for the benefit of SU treatment compared to insulin.

Many of the cases in these studies were adolescent age or younger, where the frequency of complications is expected to be low, but some individuals as old as 57 years of age were also included. The earliest report of microvascular complications occurred in the second decade of life. There was only one reported macrovascular complication in neonatal diabetes. However, most studies did not report on this complication, likely due to the overall young age of the cohort.

Recommendation: Surveillance for diabetes-related microvascular and macrovascular complications should begin in the second decade of life for permanent forms of KCNJ11-neonatal diabetes and ABCC8-neonatal diabetes, although the risk of complications occurring before the third decade of life seem low.

We did not include studies of KCNJ11 or ABCC8 carriers with transient neonatal diabetes or MODY as the duration of dysglycemia and severity of mutations is different from permanent neonatal diabetes, leading to differing diabetes prognoses; therefore, we do not provide surveillance recommendations for these phenotypes (grade D evidence).

### Recommendations for HNF1A-diabetes and HNF4A-diabetes

Diabetes onset is unusual in the first decade of life; onset is typically in the second and third decades. Later onset is also reported, although in some cases delayed recognition of diabetes is evident based on the presence of diabetes-related complications at diagnosis. Microvascular complications or risk factors for complications were present as young as 14 or 15 years of age.

There was evidence of earlier age of diagnosis in familial studies as well as an overall temporal trend in familial and larger studies of earlier diagnosis and better HbA1c. There is evidence, albeit limited, that this will translate into lower rates of diabetes-related complications and better diabetes prognoses for these forms of monogenic diabetes (Bacon 2016, Isomma). These studies also provide support for a positive impact of precision therapy with SU on diabetes-related complications but additional studies specifically designed to assess the impact of precision therapy on diabetes prognosis are needed. There is also evidence for excessive CVD risk in HNF1A-diabetes which may be directly related to pathogenic variants in*HNF1A*. The impact of statin therapy and potentially of cardioprotective glucose-lowering medications should also be a future focus of research studies.

Recommendation: Surveillance for diabetes-related microvascular and macrovascular complications should begin in the second decade of life (grade D evidence). Measures to mitigate risk of CVD should be considered in all individuals with HNF1A-diabetes even in the absence of additional traditional risk factors (grade D evidence).

### Recommendations for GCK-related hyperglycemia

GCK-related hyperglycemia uniquely seems to not have increased risk for diabetes microvascular and macrovascular complications, beyond mild retinopathy. Determining how to define the co-occurrence of type 2 diabetes, which negatively impacts prognosis, is clearly needed.

Recommendation: Surveillance for diabetes-related microvascular and macrovascular complications should begin in the fifth decade of life with earlier assessment based on symptoms or concern for co-occurrence of obesity-related insulin resistance (grade D evidence).

## Conclusions and Recommendations for Future Directions

Overall the available evidence confirms that individuals with permanent forms of KCNJ11-neonatal diabetes and ABCC8-neonatal diabetes and those with HNF1A-diabetes and HNF4A-diabetes are at high-risk for diabetes-related microvascular complications. Those with HNF1A-diabetes and HNF4A-diabetes are also at risk for macrovascular complications (with HNF1A-diabetes showing excess risk compared to unaffected relatives), but the data available for KCNJ11-neonatal diabetes and ABCC8-neonatal diabetes is limited, likely related to the relatively young age of the individuals who have been evaluated. While the impact of precision therapy is expected to improve prognosis in these forms of monogenic diabetes, the evidence to support that is limited. Retrospective, and more importantly prospective studies should focus on capturing the age of diagnosis of diabetes-related complications and assess these in the context of careful delineated details that impact prognosis, including diagnosis HbA1c, the time to achieve goal glycemic control after diagnosis, overall glycemia, diabetes duration at the start of precision treatment and glycemic response to precision versus non-precision treatment.

The overall evidence supports the low occurrence of microvascular and microvascular complications in GCK-related hyperglycemia. Future studies are needed that will focus on the outcomes of retinopathy, which are the only microvascular complication to clearly show an increased prevalence in those with GCK-related hyperglycemia. On-going surveillance of diabetes prognosis for both microvascular and macrovascular complications in older cohorts (beyond the fifth decade of life) are needed to understand the impact of lifelong mild hyperglycemia on prognosis. Additionally, future studies should attempt to identify parameters to distinguish individuals with co-occurrence of GCK-related hyperglycemia and type 2 diabetes to delineate the impact of “double-diabetes” on diabetes prognosis.

Applying a precision lens to diabetes prognosis surveillance will hopefully over time improve the diabetes-related outcomes and overall quality-of-life of those living with monogenic diabetes.

## Authors’ declaration of personal interests

RNN, CA, CV, LD, SAWG report no conflicts of interest.

## Data Availability

This is a systematic review of published, openly available studies and any additional details about the present study are available upon reasonable request to the authors

## Acknowledgements

R.N.N is supported by the following grants: ADA 7-22-ICTSPM-17; R01DK104942; U54DK118612. S.A.W.G and L.H.P. are supported by NIH NIDDK R01DK104942 and U54DK118612.

The ADA/EASD Precision Diabetes Medicine Initiative, within which this work was conducted, has received the following support: The Covidence license was funded by Lund University (Sweden) for which technical support was provided by Maria Björklund and Krister Aronsson (Faculty of Medicine Library, Lund University, Sweden). Administrative support was provided by Lund University (Malmö, Sweden), University of Chicago (IL, USA), and the American Diabetes Association (Washington D.C., USA). The Novo Nordisk Foundation (Hellerup, Denmark) provided grant support for in-person writing group meetings (PI: L Phillipson, University of Chicago, IL).

## Author contributions

*Review Design*: RNN, SAWG

*Systematic Review Implementation*: RNN, CA, LP, CV, LD, SAWG

*Data extraction manuscripts:* RNN, CA, LP, CV, LD, SAWG

*Manuscript writing:* RNN, CA, LP, CV, LD, SAWG

*Project Management:* RNN, SAWG

## References

1. Letourneau, L. R. & Greeley, S. A. W. Precision Medicine: Long-Term Treatment with Sulfonylureas in Patients with Neonatal Diabetes Due to KCNJ11 Mutations. Current diabetes reports 19, 52 (2019).

2. Schultz, C. J. et al. Microalbuminuria prevalence varies with age, sex, and puberty in children with type 1 diabetes followed from diagnosis in a longitudinal study. Oxford Regional Prospective Study Group. Diabetes Care 22, 495–502 (1999).

3. Ashcroft, F. M., Puljung, M. C. & Vedovato, N. Neonatal Diabetes and the KATP Channel: From Mutation to Therapy. Trends in endocrinology and metabolism: TEM 28, 377–387 (2017).

4. Greeley, S. A. W. et al. The cost-effectiveness of personalized genetic medicine: the case of genetic testing in neonatal diabetes. Diabetes Care 34, 622–627 (2011).

5. Gloyn, A. L. et al. Activating mutations in the gene encoding the ATP-sensitive potassium-channel subunit Kir6.2 and permanent neonatal diabetes. The New England Journal of Medicine 350, 1838–1849 (2004).

6. American Diabetes Association. 6. Glycemic Targets: Standards of Medical Care in Diabetes-2021. Diabetes Care 44, S73–S84 (2021).

7. Stride, A. et al. Cross-sectional and longitudinal studies suggest pharmacological treatment used in patients with glucokinase mutations does not alter glycaemia. Diabetologia 57, 54–56 (2014).

8. Murad, M. H., Sultan, S., Haffar, S. & Bazerbachi, F. Methodological quality and synthesis of case series and case reports. BMJ Evid Based Med 23, 60–63 (2018).

9. Cho, J. H. et al. DEND Syndrome with Heterozygous KCNJ11Mutation Successfully Treated with Sulfonylurea. Journal of Korean medical science 32, 1042–4 (2017).

10. Dupont, J. et al. Permanent neonatal diabetes mellitus due to KCNJ11 mutation in a Portuguese family: transition from insulin to oral sulfonylureas. Journal of pediatric endocrinology & metabolismfZ: JPEM 25, 367–370 (2012).

11. Gloyn, A. L. et al. KCNJ11 activating mutations are associated with developmental delay, epilepsy and neonatal diabetes syndrome and other neurological features. European journal of human geneticsfZ: EJHG 14, 824–830 (2006).

12. Heo, J. W., Kim, S.-W. & Cho, E.-H. Unsuccessful switch from insulin to sulfonylurea therapy in permanent neonatal diabetes mellitus due to an R201H mutation in the KCNJ11 gene: A case report.Diabetes Research and Clinical Practice 100, e1–e2 (2013).

13. Hindsø, M. et al. Successful Use of a GLP-1 Receptor Agonist as Add-on Therapy to Sulfonylurea in the Treatment of KCNJ11 Neonatal Diabetes. Eur J Case Rep Intern Med 8, 002352 (2021).

14. Iafusco, D. et al. No sign of proliferative retinopathy in 15 patients with permanent neonatal diabetes with a median diabetes duration of 24 years. Diabetes Care 37, e181–2 (2014).

15. John, H. et al. Neonatal diabetes is more than just a paediatric problem: 57 years of diabetes from a Kir6.2 mutation. Practical Diabetes International 22, 342–344 (2005).

16. Jones, A. G. & Hattersley, A. T. Reevaluation of a case of type 1 diabetes mellitus diagnosed before 6 months of age. Nature reviews Endocrinology 6, 347–351 (2010).

17. Klupa, T. et al. Diabetic retinopathy in permanent neonatal diabetes due to Kir6.2 gene mutations: the results of a minimum 2-year follow-up after the transfer from insulin to sulphonylurea.Diabetic medicinefZ: a journal of the British Diabetic Association 26, 663–664 (2009).

18. Klupa, T. et al. The first case report of sulfonylurea use in a woman with permanent neonatal diabetes mellitus due to KCNJ11 mutation during a high-risk pregnancy. The Journal of clinical endocrinology and metabolism 95, 3599–3604 (2010).

19. Klupa, T. et al. Efficacy and safety of sulfonylurea use in permanent neonatal diabetes due to KCNJ11 gene mutations: 34-month median follow-up. Diabetes technology & therapeutics 12, 387–391 (2010).

20. Mouler, M. et al. Clinical characteristics, growth patterns, and long-term diabetes complications of 24 patients with neonatal diabetes mellitus: A single center experience. Pediatric Diabetes 23, 45–54 (2022).

21. Stanik, J. et al. Sulfonylurea vs insulin therapy in individuals with sulfonylurea-sensitive permanent neonatal diabetes mellitus, attributable to a KCNJ11 mutation, and poor glycaemic control.Diabetic Medicine 35, 386–391 (2018).

22. Tammaro, P. et al. A Kir6.2 mutation causing severe functional effects in vitro produces neonatal diabetes without the expected neurological complications. Diabetologia 51, 802–810 (2008).

23. Bowman, P. et al. Effectiveness and safety of long-term treatment with sulfonylureas in patients with neonatal diabetes due to KCNJ11 mutations: an international cohort study.The Lancet Diabetes & Endocrinology 6, 637–646 (2018).

24. Thurber, B. W. et al. Age at the time of sulfonylurea initiation influences treatment outcomes in KCNJ11-related neonatal diabetes. Diabetologia 58, 1430–1435 (2015).

25. Stankute, I. et al. Systematic Genetic Study of Youth With Diabetes in a Single Country Reveals the Prevalence of Diabetes Subtypes, Novel Candidate Genes, and Response to Precision Therapy.Diabetes 69, 1065–1071 (2020).

26. Warncke, K. et al. Clinical presentation and long-term outcome of patients with KCNJ11/ABCC8 variants: Neonatal diabetes or MODY in the DPV registry from Germany and Austria. Pediatric Diabetes 23, 999– 1008 (2022).

27. Bowman, P. et al. Long-term Follow-up of Glycemic and Neurological Outcomes in an International Series of Patients With Sulfonylurea-Treated ABCC8 Permanent Neonatal Diabetes.Diabetes Care 44, 35–42 (2021).

28. Esmatjes, E. et al. Neonatal diabetes with end-stage nephropathy: pancreas transplantation decision. Diabetes Care 31, 2116–2117 (2008).

29. Docena, M. K., Faiman, C., Stanley, C. M. & Pantalone, K. M. Mody-3: Novel HNF1A mutation and the utility of glucagon-like peptide (GLP)-1 receptor agonist therapy. Endocrine Practice 20, 107–111 (2014).

30. Dusatkova, P. et al. HNF1A mutation presenting with fetal macrosomia and hypoglycemia in childhood prior to onset of overt diabetes. Journal of pediatric endocrinology & metabolismfZ: JPEM 24, 187–189 (2011).

31. Egan, A. M., Cunningham, A., Jafar-Mohammadi, B. & Dunne, F. P. Diabetic ketoacidosis in the setting of HNF1A-maturity onset diabetes of the young. BMJ Case Reports 2015, (2015).

32. Hummel, M. et al. Two Caucasian families with the hepatocyte nuclear factor-1alpha mutation Tyr218Cys. Experimental and Clinical Endocrinology and Diabetes 115, 62–64 (2007).

33. Malecki, M. T. et al. Renal malformations may be linked to mutations in the hepatocyte nuclear factor-1α (MODY3) gene. Diabetes Care 28, 2774–2776 (2005).

34. Menzel, R. et al. A low renal threshold for glucose in diabetic patients with a mutation in the hepatocyte nuclear factor-1alpha (HNF-1alpha) gene. Diabet Med 15, 816–20 (1998).

35. Ng, M. C. et al. Familial early-onset type 2 diabetes in Chinese patients: obesity and genetics have more significant roles than autoimmunity. Diabetes Care 24, 663–71 (2001).

36. Ovsyannikova, A. K. et al. A Case of Maturity Onset Diabetes of the Young (MODY3) in a Family with a Novel HNF1A Gene Mutation in Five Generations. Diabetes Therapy 9, 413–420 (2018).

37. Ren, X. Y. et al. Clinical Characteristics and Gene Mutations of Two Families with MODY 3 in Inner Mongolia. Pharmacogenomics and Personalized Medicine 15, 1019–1027 (2022).

38. Saudek, F. et al. Maturity-onset diabetes of the young with end-stage nephropathy: a new indication for simultaneous pancreas and kidney transplantation? Transplantation 77, 1298–1301 (2004).

39. To, C., Liu, L., Satoskar, R. S. & Thuluvath, P. J. Diabetic Hepatosclerosis in a Woman with Maturity-Onset Diabetes of the Young Type 3. Digestive Diseases and Sciences 67, 2688–2690 (2022).

40. Weinert, L. S. et al. Three unreported glucokinase (GCK) missense mutations detected in the screening of thirty-two Brazilian kindreds for GCK and HNF1A-MODY. Diabetes Research and Clinical Practice 106, e44– e48 (2014).

41. Willson, J. S., Godwin, T. D., Wiggins, G. A., Guilford, P. J. & McCall, J. L. Primary hepatocellular neoplasms in a MODY3 family with a novel HNF1A germline mutation. J Hepatol 59, 904–7 (2013).

42. Zhang, M., Wang, T., Shi, L. & Yang, Y. Hepatocyte nuclear factor-α genetic mutation in a Chinese pedigree with maturity-onset diabetes of the young (MODY3). Diabetes/Metabolism Research and Reviews 31, 767– 770 (2015).

43. Bacon, S. et al. Successful maintenance on sulphonylurea therapy and low diabetes complication rates in a HNF1A-MODY cohort. Diabet Med 33, 976–984 (2016).

44. Bacon, S. et al. Circulating CD36 is reduced in HNF1A-MODY carriers. PLoS One 8, e74577 (2013).

45. Barrio, R. et al. Nine novel mutations in maturity-onset diabetes of the young (MODY) candidate genes in 22 Spanish families. The Journal of clinical endocrinology and metabolism 87, 2532–2539 (2002).

46. Isomaa, B. et al. Chronic diabetic complications in patients with MODY3 diabetes. Diabetologia vol. 41 467–73 (1998).

47. Iwasaki, N. et al. Liver and kidney function in Japanese patients with maturity-onset diabetes of the young. Diabetes Care 21, 2144–8 (1998).

48. Lebenthal, Y. et al. The unique clinical spectrum of maturity onset diabetes of the young type 3. Diabetes Research and Clinical Practice 135, 18–22 (2018).

49. Lee, B. C., Appleton, M., Shore, A. C., Tooke, J. E. & Hattersley, A. T. Impaired maximum microvascular hyperaemia in patients with MODY 3 (hepatocyte nuclear factor-1alpha gene mutations).Diabetic medicinefZ: a journal of the British Diabetic Association 16, 731–735 (1999).

50. Ma, Y. et al. New clinical screening strategy to distinguish HNF1A variant-induced diabetes from young early-onset type 2 diabetes in a Chinese population. BMJ Open Diabetes Res Care 8, (2020).

51. Poitou, C. et al. Maturity onset diabetes of the young: clinical characteristics and outcome after kidney and pancreas transplantation in MODY3 and RCAD patients: a single center experience.Transpl Int 25, 564–72 (2012).

52. Sagen, J. V., Njølstad, P. R. & Søvik, O. Reduced prevalence of late-diabetic complications in MODY3 with early diagnosis [1]. Diabetic Medicine 19, 697–698 (2002).

53. Schober, E. et al. Phenotypical aspects of maturity-onset diabetes of the young (MODY diabetes) in comparison with Type 2 diabetes mellitus (T2DM) in children and adolescents: experience from a large multicentre database. Diabetic medicinefZ: a journal of the British Diabetic Association 26, 466–473 (2009).

54. Skupien, J. et al. Molecular background and clinical characteristics of HNF1A MODY in a Polish population. Diabetes Metab 34, 524–8 (2008).

55. Steele, A. M. et al. Increased all-cause and cardiovascular mortality in monogenic diabetes as a result of mutations in the HNF1A gene. Diabetic medicinefZ: a journal of the British Diabetic Association 27, 157–161 (2010).

56. Szopa, M. et al. A decision algorithm to identify patients with high probability of monogenic diabetes due to HNF1A mutations. Endocrine 1–7 (2019) doi:10.1007/s12020-019-01863-7.

57. Szopa, M. et al. Quality of life assessment in patients with HNF1A-MODY and GCK-MODY. Endocrine 64, 246–253 (2019).

58. Szopa, M. et al. Prevalence of Retinopathy in Adult Patients with GCK-MODY and HNF1A-MODY. Experimental and Clinical Endocrinology & Diabetes 123, 524–528 (2015).

59. Szopa, M. et al. Intima-media thickness and endothelial dysfunction in GCK and HNF1A-MODY patients. Eur J Endocrinol 172, 277–83 (2015).

60. Velho, G., Vaxillaire, M., Boccio, V., Charpentier, G. & Froguel, P. Diabetes complications in NIDDM kindreds linked to the MODY3 locus on chromosome 12q. Diabetes Care 19, 915–9 (1996).

61. Xu, J. Y. et al. Genetic and clinical characteristics of maturity-onset diabetes of the young in Chinese patients. European Journal of Human Genetics 13, 422–427 (2005).

62. Hara, K. et al. Maturity-onset diabetes of the young resulting from a novel mutation in the HNF-4alpha gene. Intern Med 41, 848–52 (2002).

63. Lindner, T. et al. Hepatic function in a family with a nonsense mutation (R154X) in the hepatocyte nuclear factor-4alpha/MODY1 gene. The Journal of clinical investigation 100, 1400–1405 (1997).

64. Spiro, A. J., Vu, K. N. & Warnock, A. L. An Atypical HNF4A Mutation Which Does Not Conform to the Classic Presentation of HNF4A-MODY. Case Reports in Endocrinology 2018, (2018).

65. Berger, M. et al. Are glucokinase mutations associated with low triglycerides?Clinical Chemistry 51, 791– 793 (2005).

66. Celikay, O. et al. Ocular surface assessment in maturity-onset diabetes of the young. International Journal of Diabetes in Developing Countries 41, 136–140 (2021).

67. Franco, L. F. et al. Cardiovascular risk assessment by coronary artery calcium score in subjects with maturity-onset diabetes of the young caused by glucokinase mutations.Diabetes Res Clin Pract 176, 108867 (2021).

68. Gjesing, A. P. et al. 14-fold increased prevalence of rare glucokinase gene variant carriers in unselected Danish patients with newly diagnosed type 2 diabetes. Diabetes Res Clin Pract 194, 110159 (2022).

69. Kawakita, R. et al. Molecular and clinical characterization of glucokinase maturity-onset diabetes of the young (GCK-MODY) in Japanese patients. Diabetic medicinefZ: a journal of the British Diabetic Association (2014) doi:10.1111/dme.12487.

70. Njølstad, P. R., Cockburn, B. N., Bell, G. I. & Søvik, O. A missense mutation, Val62Ala, in the glucokinase gene in a Norwegian family with maturity-onset diabetes of the young. Acta Paediatrica, International Journal of Paediatrics 87, 853–856 (1998).

71. Page, R. C. et al. Clinical characteristics of subjects with a missense mutation in glucokinase.Diabetic medicinefZ: a journal of the British Diabetic Association 12, 209–217 (1995).

72. Pruhova, S. et al. Chronic mild hyperglycemia in GCK-MODY patients does not increase carotid intima-media thickness. International Journal of Endocrinology 2013, (2013).

73. Sagen, J. V. et al. Diagnostic screening of MODY2/GCK mutations in the Norwegian MODY Registry. Pediatric Diabetes 9, 442–449 (2008).

74. Steele, A. M. et al. Prevalence of vascular complications among patients with glucokinase mutations and prolonged, mild hyperglycemia. JAMA: The Journal of the American Medical Association 311, 279–286 (2014).

75. Velho, G. et al. Identification of 14 new glucokinase mutations and description of the clinical profile of 42 MODY-2 families. Diabetologia vol. 40 217–24 (1997).

76. Amin, R. et al. Risk of microalbuminuria and progression to macroalbuminuria in a cohort with childhood onset type 1 diabetes: prospective observational study. BMJ 336, 697–701 (2008).

