## Supplemental Table 1 for "Systematic Review of Monogenic Diabetes Prognostics"

**Supplemental Table 1:** Search terms and keywords used to identify relevant studies for GCK-related hyperglycemia, HNF1A-diabetes, HNF4A-diabetes

| Diabetes phenotypes | Genes* | Treatment Class** | Glycemic measure | Microvascular Outcomes | Macrovascular Outcomes |
|---|---|---|---|---|---|
| Monogenic diabetes<br>MODY<br>Maturity onset diabetes of the young<br>Maturity-onset diabetes of the young<br>Mature onset diabetes of the young | HNF1A<br>HNF4A<br>GCK | Metformin<br>Sulphonylureas<br>Glinides<br>Thiazolidinediones<br>Alpha glucosidase inhibitors<br>DPP-4 inhibitors<br>SGLT2 inhibitors<br>GLP-1 R Agonists<br>Insulin | HbA1c<br>Hemoglobin A1c<br>Glycated hemoglobin<br>Glycemia<br>Treatment response | Nephropathy<br>Proteinuria<br>Macroalbuminuria<br>Renal impairment<br>Chronic kidney disease<br>Microalbuminuria<br>End-stage renal disease<br>ESRD<br>Kidney failure | Cardiovascular disease<br>Myocardial infarction<br>Revascularization<br>Acute coronary syndrome Coronary heart disease<br>Heart failure<br>Cardiac Failure<br>Angioplasty<br>Percutaneous Coronorary Intervention<br>Cardiovascular Mortality MACE<br>Stroke<br>TIA |
| Neonatal diabetes<br>PNDM<br>Infancy-onset diabetes<br>NDM<br>TNDM<br>Permanent neonatal diabetes<br>Transient neonatal diabetes | KCNJ11<br>ABCC8 | | | Retinopathy | |
| | | | | Neuropathy | |

| **Genes*** | | | | |
|---|---|---|---|---|
| HNF1A | HNF4A | GCK | KCNJ11 | ABCC8 |
| Hepatocyte Nuclear Factor 1 Alpha | Hepatocyte Nuclear Factor 4 Alpha | Glucokinase | Potassium Inwardly Rectifying Channel Subfamily J Member 11 | ATP Binding Cassette Subfamily C Member 8 |
| HNF1 | HNF4 | HK4 | ATP-Sensitive Inward Rectifier Potassium Channel 11 | SUR1 |
| Hepatocyte Nuclear Factor 1-Alpha | Hepatocyte Nuclear Factor 4-Alpha | Glucokinase (Hexokinase 4) | BIR | ABC36 |
| Transcription Factor HNF-1 | Transcription Factor HNF-4 | Hexokinase Type IV | Potassium Channel, Inwardly Rectifying Subfamily J Member 11 | TNDM2 |
| Hepatic Nuclear Factor 1 Alpha | Hepatic Nuclear Factor 4 Alpha | Hexokinase-4 | Inward Rectifier K(+) Channel Kir6.2 | HRINS |
| HNF1alpha | HNF4alpha | HK IV | Kir6.2 | PHHI |
| HNF-1-Alpha | HNF-4-Alpha | Hexokinase D, Pancreatic Isozyme | IKATP | MRP8 |
| HNF1alpha | HNF4alpha | Hexokinase 4 | Potassium Inwardly-Rectifying Channel, Subfamily J, Member 11 | HHF1 |
| HNF1a | HNF4a | Hexokinase-D | Inwardly-Rectifying Potassium Channel Subfamily J Member 11 | SUR |
| MODY3 | MODY1 | MODY2 | Inwardly Rectifing Potassium Channel Subfamily J Member 11 | HI |
| HNF1A | HNF4A | GCK | Potassium Channel Inwardly Rectifing Subfamily J Member 11 | ATP-Binding Cassette, Sub-Family C (CFTR/MRP), Member 8 |
| TCF-1 | TCF-14 | MODY-2 | Potassium Voltage-Gated Channel Subfamily J Member 11 | ATP-Binding Cassette Sub-Family C Member 8 |
| TCF1 | TCF14 | | Inwardly Rectifying Potassium Channel KIR6.2 | Sulfonylurea Receptor (Hyperinsulinemia) |

| | | | | |
|---|---|---|---|---|
| MODY-3 | MODY-1 | | Beta-Cell Inward Rectifier Subunit | Sulfonylurea Receptor 1 |
| | | | Beta-Cell Inward Rectifier | ATP-Binding Cassette Transporter Sub-Family C Member 8 |
| | | | KIR6.2 | SUR1delta2 |
| | | | PNDM2 | PNDM3 |
| | | | HHF2 | Sulphonylurea Receptor 1 |
| | | | PHHI | SUR1delta2KATP |
| | | | TNDM3 | ATP binding cassette subfamily C member 8 |
| | | | Katp-channel | |
| | | | Katp | |
| | | | KATP | |

| **Treatment Class**** | | | | | | | | |
|---|---|---|---|---|---|---|---|---|
| Metformin | Sulphonylureas | Glinides | Thiazolidine-diones | Alpha glucosidase inhibitors | DPP-4 inhibitors | SGLT2 inhibitors | GLP-1 R Agonists | Insulin |
| Metformin<br>Biguanide | Sulphonylurea<br>Sulfonylurea<br>Gliclazide<br>Glipizide<br>Glibenclamide<br>Glyburide<br>Glimepiride<br>Tolbutamide<br>Chlorpro-pamide | Repaglinide<br>Nateglinide | Thiazolidine-dione<br>PPARg Agonist<br>Rosiglitazone<br>Pioglitazone<br>Troglitazone | Alphagluco-sidase inhibitor<br>Acarbose | DPP4 inhibitor<br>DPP-4 inhibitor<br>Dipeptidyl-peptidase 4 inhibitor<br>Dipeptidyl-peptidase-4 inhibitor<br>Sitagliptin<br>Vildagliptin<br>Saxagliptin<br>Linagliptin<br>Alogliptin | SGLT2 inhibitor<br>SGLT2i<br>SGLT-2 inhibitor<br>SGLT-2i<br>Sodium Glucose Transporter 2 inhibitor<br>Dapagliflozin<br>Empagliflozin<br>Ertagliflozin<br>Canagliflozin | GLP1RA<br>GLP1 Receptor Agonists<br>GLP-1 Receptor Agonists<br>GLP-1RA<br>Exenatide<br>Liraglutide<br>Lixisenatide<br>Semaglutide<br>Dulaglutide<br>Albiglutide | Isophane<br>NPH insulin<br>Basal Insulin<br>Long acting insulin<br>Glargine<br>Detemir<br>Degludec<br>Insulin |
